# Polygenic associations with clinical and neuropathological trait heterogeneity across TDP-43 proteinopathies

**DOI:** 10.1101/2023.10.05.23296613

**Authors:** Barbara E. Spencer, David J. Irwin, Vivianna M. Van Deerlin, EunRan Suh, Edward B. Lee, Lauren Elman, Colin C. Quinn, Defne Amado, Michael Baer, Murray Grossman, David A. Wolk, Corey T. McMillan

## Abstract

**Objective:** TDP-43 proteinopathies, including amyotrophic lateral sclerosis (ALS), frontotemporal lobar degeneration with TDP-43 (FTLD-TDP), and limbic-predominant age-related TDP-43 encephalopathy, encompass a spectrum of clinical and neuropathological traits. Despite mounting evidence for shared genetic risk across TDP-43 proteinopathies, the modifiers of individual-level traits are unknown. We aimed to identify polygenic contributions to trait heterogeneity across TDP-43 proteinopathies.

**Methods:** We used weighted correlation analysis of GWAS summary statistics for ALS, FTLD-TDP, and hippocampal sclerosis of aging (HS-Aging) to identify data-driven clusters of highly correlated single nucleotide polymorphisms (SNPs). We performed gene ontology enrichment analysis for each identified cluster. We derived cluster-specific polygenic scores and evaluated their association with clinical and neuropathological traits in an independently evaluated sample of individuals who met neuropathological and/or genetic criteria for FTLD-TDP or ALS (n=260).

**Results:** We identified 5 distinct data-driven clusters, including 3 GWAS phenotype-specific clusters (FTLD-TDP, ALS, HS-Aging) and 2 clusters representing the overlap between a pair of GWAS phenotypes (ALS-FTLD and FTLD-HS). Pathway analysis revealed biologically meaningful associations including distinct GWAS phenotype-specific processes within clusters. Cluster-specific ALS and FTLD-TDP polygenic risk each associated with individual-level clinical traits, even within the context of autosomal dominant mutation carriers, where higher ALS polygenic risk associated with neuromuscular impairment and higher FTLD-TDP polygenic risk associated with cognitive-behavioral impairment. Moreover, higher FTLD-TDP polygenic risk associated with higher TDP-43 burden within characteristic FTLD-TDP brain regions.

**Interpretation:** We suggest that there are polygenic modifiers of clinical and neuropathological traits across TDP-43 proteinopathies that may contribute to individual-level differences, including likelihood for developing FTLD or ALS.

## INTRODUCTION

TAR DNA-binding protein ∼43kDa (TDP-43) inclusions are the pathological hallmark of frontotemporal lobar degeneration with TDP-43 (FTLD-TDP), >98% of amyotrophic lateral sclerosis (ALS), and ∼90% of hippocampal sclerosis of aging (HS-Aging).^4–7^ Despite shared neuropathological traits, individuals with TDP-43 proteinopathies present with heterogeneous clinical traits including neuromuscular impairments (i.e., ALS) and cognitive-behavioral impairments (i.e., FTLD-TDP). TDP-43 pathology and neurodegeneration are observed in characteristic neuroanatomical regions that correlate with clinical traits. Some individuals present with both neuromuscular and cognitive-behavioral impairment (i.e., ALS-FTD) which, together with the shared neuropathological traits support the notion that FTLD-TDP and ALS are part of a clinicopathologic spectrum.^8^ Alternatively, individuals with HS-Aging typically present with an amnestic syndrome with an older age of onset.^9–11^ Many efforts have focused on the identification of genetic variants that predispose individuals to FTLD-TDP, ALS, or HS-Aging. However, the contribution of genotypes to the shared and disparate clinical and neuropathologic traits observed across TDP-43 proteinopathies is unclear.

Large scale case-control genome wide association studies (GWAS) have focused on the identification of single nucleotide polymorphisms (SNPs) associated with risk for ALS,^1,12^ FTLD-TDP,^2^ or HS-Aging.^3,6^ Several large-scale GWAS established ALS as a complex, polygenic disease.^1,12,13^ The first GWAS in FTLD-TDP cases identified three SNPs spanning *TMEM106B* that reached genome-wide significance and several studies have validated the importance of these loci in FTLD pathogenesis.^14–19^ GWAS of HS-Aging have also implicated several SNPs, including those in *TMEM106B.*^3,6^ Additional efforts have aimed to uncover a shared genetic basis to explain the biological mechanisms that drive susceptibility in FTLD-TDP and ALS. A meta-analysis that jointly analyzed ALS and FTLD-TDP cases identified significant associations in *UNC13A*^20^ which had previously only been identified as a susceptibility SNP for ALS and has since been implicated in an FTLD-TDP GWAS.^21^ Another study used a genetic enrichment approach that found an enrichment of ALS as a function of increased association with FTLD-TDP, indicating these phenotypes are genetically related.^22^ However, conjunction analyses performed to identify which SNPs were jointly associated with increased risk for ALS and FTLD-TDP only revealed a SNP that is used as a surrogate marker for *C9orf72* repeat expansions, which can cause FLTD-TDP and/or ALS even within the same family.^23,24^

Despite mounting evidence for shared genetic risk, considerably less effort has focused on the modifiers of individual-level traits. Rather than focusing only on overlapping risk factors, identifying disparate risk alleles may reveal specific neuroanatomic and/or cellular vulnerabilities that contribute to our understanding of trait-specific drivers. Using existing GWAS summary statistics with novel bioinformatics strategies, we aimed to identify polygenic contributions to individual-level clinical and neuropathological traits.

We compared the relative associations of case-control GWAS summary statistics for ALS, FTLD-TDP, and HS-aging to identify data-driven clusters of highly correlated SNPs that may contribute to clinical and neuropathological trait heterogeneity across these neurodegenerative diseases. We identified the biological pathways associated with these clusters. We then related these cluster SNPs to individual-level clinical and neuropathological traits within a neuropathologically and/or genetically defined sample of ALS and FTLD-TDP cases, including the presence of neuromuscular impairment and/or cognitive-behavioral impairment and the burden of TDP-43 pathology across brain regions, in a well-characterized sample of more than 250 individuals with ALS and/or FTLD-TDP, through the construction of cluster-specific polygenic risk scores (PRS). We identified clusters of genotypic contributions to selective vulnerability in the anatomic distribution of TDP-43 associated disease, and, for the first time, established potential genetic contributions to risk for either neuromuscular or cognitive-behavioral impairments within individuals with *C9orf72* repeat expansions.

## SUBJECTS/MATERIALS AND METHODS

## Cluster identification and annotation

### Summary Statistic Data

To derive clusters of associated SNPs, we performed weighted correlation analysis on publicly-available summary statistics for reported case-control ALS,^1^ FTLD-TDP,^2^ and HS-Aging^3^ (which is enriched for TDP-43^5–7^) GWAS. The ALS GWAS included a combined discovery sample of 20,806 sporadic and familial ALS cases. 12,577 cases were included from summary statistics downloaded from van Rheenen et al^13^. The remaining 8,229 cases were of non-Hispanic white race/ethnicity and were diagnosed with ALS according to the El Escorial criteria^25^. The FTLD-TDP GWAS included 515 FTLD-TDP individuals of European descent with dementia (with or without MND) and either a pathological diagnosis of FTLD-TDP (n=499) or, in the case of living individuals, a pathogenic GRN mutation (n=16). The HS-Aging GWAS evaluated 310 cases of HS-Aging as defined by presence vs. absence at pathological evaluation.

To harmonize summary statistics across GWAS, we only evaluated SNPs that were commonly genotyped across GWAS. FTLD-TDP summary statistics were transformed from NCBI36/hg18 into GRCh37/hg19 using liftOver^26^ for comparison with the ALS and HS-Aging summary statistics. Following alignment, we identified 494,417 SNPs commonly genotyped across these GWAS summary statistics. GWAS statistics for each of the three studies were transformed into z-scores to reduce the effects of sample size differences, and only the absolute value of the z-score was used for analysis. For the ALS GWAS, z-scores were calculated as the effect divided by the standard error. For the FTLD-TDP and HS-Aging GWAS, z-scores were calculated as |z|=|Φ−1(p/2)| where Φ−1 is the inverse cumulative distribution function of the normal distribution, and p is the p-value. The top 1% of SNPs with the strongest associations to any GWAS phenotype (by |z|, n= 4,945) were selected for analysis (Fig. S1).

### Weighted Correlation Analysis

The WGCNA package in R^27^ was used to perform weighted correlation analysis. This approach uses the patterns of associations of SNPs with the respective GWAS phenotypes to identify correlations between SNPs and construct clusters based on those correlations. First, an adjacency matrix is constructed using the strength of the association between each SNP and each GWAS phenotype. From the adjacency matrix values, a topological overlap matrix is created to alleviate the effect of noise, and this is converted into a distance matrix which is used as the basis for clustering. Clusters are identified with unsupervised clustering (i.e., without the use of a priori defined gene or SNP sets), specifically hierarchical clustering, along with dynamic tree cut.

As only SNPs that were common across GWAS were considered for analysis, there were 0 SNPs removed due to missingness. Clusters were constructed with a minimum size of 30 SNPs. Clusters whose distance was less than 0.25 were merged. To test for robustness, we created 100 samples, each containing a random 80% of the SNPs selected for weighted correlation analysis, without replacement (Fig. S2). For each resample, weighted correlation analysis was performed as in the full sample.

After clusters were identified, multialleleic SNPs and SNPs with > 10% sample missingness in the individual level genotype data (see Genotype Data) were removed, and SNPs were pruned within clusters using an R^2^ threshold of 0.3 with LDlinkR.^28^

### Cluster Annotation

To annotate, cluster SNPs were mapped to overlapping genes using the biomaRt package in R^29^, specifically ENSEMBL_MART_SNP. A gene ontology enrichment analysis for biological processes was performed separately for each cluster using gprofiler2. ^30^

Enrichment was determined by overrepresentation test, which calculates an expected value by matching the proportion of genes in a particular annotation data category in the human genome reference list to the number of genes provided and calculates a fold enrichment value by comparing the true number of genes in a particular annotation data category to the expected value. A p-value was calculated for each term with the hypergeometric test followed by correction for multiple testing using the False Discovery Rate (FDR), calculated by the Benjamini-Hochberg procedure. Pathways that met FDR p<.05 were examined for each cluster.

## Individual-level data in a neuropathologically and/or genetically defined sample

### Case selection

To ensure a TDP-43 proteinopathy, we selected 260 individuals who met neuropathological criteria for FTLD-TDP (n=75) and/or ALS (n=105) or had a pathogenic mutation associated with TDP-43 (n=135, including those who met neuropathological criteria) to include for analysis, described in detail below. *SOD1* ALS cases and *VCP* tauopathy cases were excluded to focus on TDP-43 proteinopathy.

### Genotype Data

Individual level imputed genotype data, derived from a variety of prior GWAS array genotyping chips, was available in the Penn Integrated Neurodegenerative Disease Database^31^. To harmonize data, we performed imputation of allele dosages on samples passing QC using the Michigan Imputation Server^32^ with the TOPMed reference panel based on >97K sequenced samples.^33^ Prior to genotype analyses or imputation, all genotype data were QCed using standard metrics including but not limited to sex discrepancy, sample relatedness, heterozygosity, and Hardy-Weinberg equilibrium.^34^

### Genetic Sequencing

DNA was extracted from peripheral blood or frozen brain tissue following the manufacturer’s protocols (QuickGene DNA whole blood kit (Autogen) for blood, and QIAamp DNA Mini Kit (Qiagen) for brain tissue). All individuals were tested for *C9orf72* hexanucleotide repeat expansions using a modified repeat-primed PCR as previously described ^35^ and screened for mutations associated with FTD and/or ALS using whole exome/genome sequencing, and/or a custom targeted multi neurodegenerative disease sequencing panel ^36^ which included *SOD1*^37^, *TBK1*^38^, *TARDBP* ^39^, *VCP* ^40^, and *GRN*^41,42^). The sequencing data was analyzed using Geneticist Assistant software (Soft Genetics, State College, PA).

### Polygenic risk score calculation

The weighted correlation analysis identified 5 clusters of correlated SNPs (see Data-driven clusters of correlated SNPs identified using summary statistics). We limited our analysis to clusters associated with a single GWAS phenotype (C1 ALS, C2 FTLD-TDP, and C3 HS-Aging) in an effort to focus on GWAS phenotype-specific drivers. Due to a lack of directionality information in the provided summary statistics, we were unable to calculate a PRS for C3 HS-Aging.

Cluster-specific PRS were constructed for C1 ALS and C2 FTLD-TDP as the weighted sum of alleles per SNP per cluster for each individual, using z-score transformed associations with the respective GWAS phenotype, ALS or FTLD-TDP. The effect allele was labelled as ‘Allele1’ in the ALS summary statistics and was the minor allele in the FTLD-TDP summary statistics. Here, we assigned minor allele using the European 1,000 Genomes sample allele frequencies. The direction of the weight for each SNP was determined by whether the dose of the effect (direction consistent with summary statistic) or non-effect (direction flipped) allele was provided in the individual-level data.

Each PRS was centered using the European 1,000 Genomes sample allele frequencies and mean scaled. Missing allele dosages were replaced with the European 1,000 Genomes sample average. SNPs without allele frequency information were removed. The C1 ALS PRS included 1021 SNPs and the C2 FTLD-TDP PRS included 1013 SNPs. Individuals with >10% SNP missingness for either PRS were removed. This resulted in the removal of 130 individuals.

PRS were tested for association with clinical and neuropathological traits using one-way ANOVAs controlling for age at last contact and sex. Some individuals in the sample were included in a prior ALS or FTLD-TDP GWAS for which summary statistics were used to generate the clusters (Table 1). Therefore, inclusion in a prior ALS or FTLD-TDP GWAS was additionally controlled for in all ANOVAs.

**Table 1.** Sample Characteristics.

|  | MND-only | Cog+MND | Cog-only |
| --- | --- | --- | --- |
| **n** | 125 | 31 | 104 |
| **Sex (Male, %)** | 72 (57.6) | 17 (54.8) | 59 (56.7) |
| **Education (mean (SD))** | 14.88 (3.09) | 14.43 (2.66) | 15.67 (3.36) |
| **Deceased (%)** | 121 (96.8) | 31 (100.0) | 86 (82.7) |
| **Age at last contact (mean (SD))** | 62.16 (10.21) | 60.42 (8.55) | 68.54 (8.47) |
| **Race (%)** |  |  |  |
| *Asian* | 0 (0.0) | 0 (0.0) | 1 (1.0) |
| *Black or African American* | 3 (2.4) | 1 (3.2) | 1 (1.0) |
| *More than one race* | 0 (0.0) | 0 (0.0) | 1 (1.0) |
| *White* | 121 (96.8) | 30 (96.8) | 99 (95.2) |
| *Unknown or not reported* | 1 (0.8) | 0 (0.0) | 2 (1.9) |
| **Ethnicity (%)** |  |  |  |
| *Hispanic or Latino* | 2 (1.6) | 0 (0.0) | 1 (1.0) |
| *Not Hispanic or Latino* | 120 (96.0) | 31 (100.0) | 101 (97.1) |
| *Unknown or not reported* | 3 (2.4) | 0 (0.0) | 2 (1.9) |
| **Mutation (%)** |  |  |  |
| *No known mutation* | 77 (61.6) | 7 (22.6) | 41 (39.4) |
| *C9orf72* | 47 (37.6) | 24 (77.4) | 42 (40.4) |
| *GRN* | 0 (0.0) | 0 (0.0) | 17 (16.3) |
| *TARDBP* | 1 (0.8) | 0 (0.0) | 0 (0.0) |
| *TBK1* | 0 (0.0) | 0 (0.0) | 2 (1.9) |
| *VCP* | 0 (0.0) | 0 (0.0) | 2 (1.9) |
| **Prior GWAS Case\* (%)** |  |  |  |
| *No prior GWAS inclusion* | 64 (51.2) | 15 (48.4) | 82 (78.8) |
| *ALS GWAS* | 61 (48.8) | 12 (38.7) | 0 (0.0) |
| *FTLD-TDP GWAS* | 0 (0.0) | 4 (12.9) | 22 (21.2) |
| **TDP Type (%)** |  |  |  |
| *A* | 4 (4.4) | 2 (11.8) | 26 (39.4) |
| *B* | 6 (6.6) | 9 (52.9) | 12 (18.2) |
| *C* | 0 (0.0) | 0 (0.0) | 23 (34.8) |
| *D* | 0 (0.0) | 0 (0.0) | 2 (3.0) |
| *E* | 5 (5.5) | 1 (5.9) | 2 (3.0) |
| *Not specified* | 76 (83.5) | 5 (29.4) | 1 (1.5) |

Demographic, genetic, and pathological data for individuals with neuromuscular impairment (MND-only), cognitive-behavioral impairment (Cog-only), or any evidence of a combination of impairment (Cog+MND) within the sample.
\*Inclusion in either the ALS<sup>1</sup> or FTLD-TDP<sup>2</sup> GWAS for which summary statistics were used to generate the clusters.

### Clinical Traits

Within known TDP-43 proteinopathy cases, individuals were characterized based on clinical traits of neuromuscular impairment (MND-only), cognitive-behavioral impairment (Cog-only), and any evidence of a combination of impairment (Cog+MND) at any point in the natural history of disease determined through clinical and research chart review.

### Neuropathological Traits

Detailed neuropathological assessments were performed using established and uniform methods of fixation, tissue processing, IHC with well-characterized antibodies, and current neuropathological criteria, as has been described in detail elsewhere.^36,43,44^ Briefly, 15 brain regions from one hemisphere, alternating right and left at random, are routinely sampled at autopsy, formalin-fixed, and processed for immunohistochemical staining using 1D3 for phosphorylated TDP-43. Each brain region was semi-quantitatively scored for the burden of TDP-43 (0, absent; 0.5, rare; 1, mild; 2, moderate; 3, severe). ALS was diagnosed based on the presence of upper and lower motor neuron degeneration, and TDP-43 subtypes were determined based on TDP-43 immunohistochemistry as previously described.^45^

## RESULTS

### Data-driven clusters of correlated SNPs identified using summary statistics

Weighted correlation network analysis identified 5 clusters of correlated SNPs associated with either a single GWAS phenotype (accordingly referred to as C1 ALS, C2 FTLD-TDP, and C3 HS-Aging from now on) or a pair of GWAS phenotypes (accordingly referred to as C4 ALS-FTLD and C5 FTLD-HS from now on, Fig. 1). Cluster assignment was robust, particularly for SNPs associated with a single GWAS phenotype where >97% of SNPs were assigned to the same cluster (C1 ALS, C2 FTLD-TDP, or C3 HS-Aging) when weighted correlation analysis was performed 100 times, each time including a random 80% of the 4,945 SNPs (Fig. S2). After clusters were identified, SNPs were LD pruned within clusters to reduce redundancy. After pruning, C1 ALS contained 1,025 SNPs, C2 FTLD-TDP contained 1,021 SNPs, C3 HS-Aging contained 646 SNPs, C4 ALS-FTLD contained 80 SNPs, and FTLD-HS contained 72 SNPs (Fig. S3). In the remaining sections we identify the biological pathways associated with these cluster SNPs and test for individual-level associations between cluster-specific polygenic risk and clinical and neuropathological traits of individuals who met neuropathological and/or genetic criteria for FTLD-TDP or ALS.

**Fig. 1.**
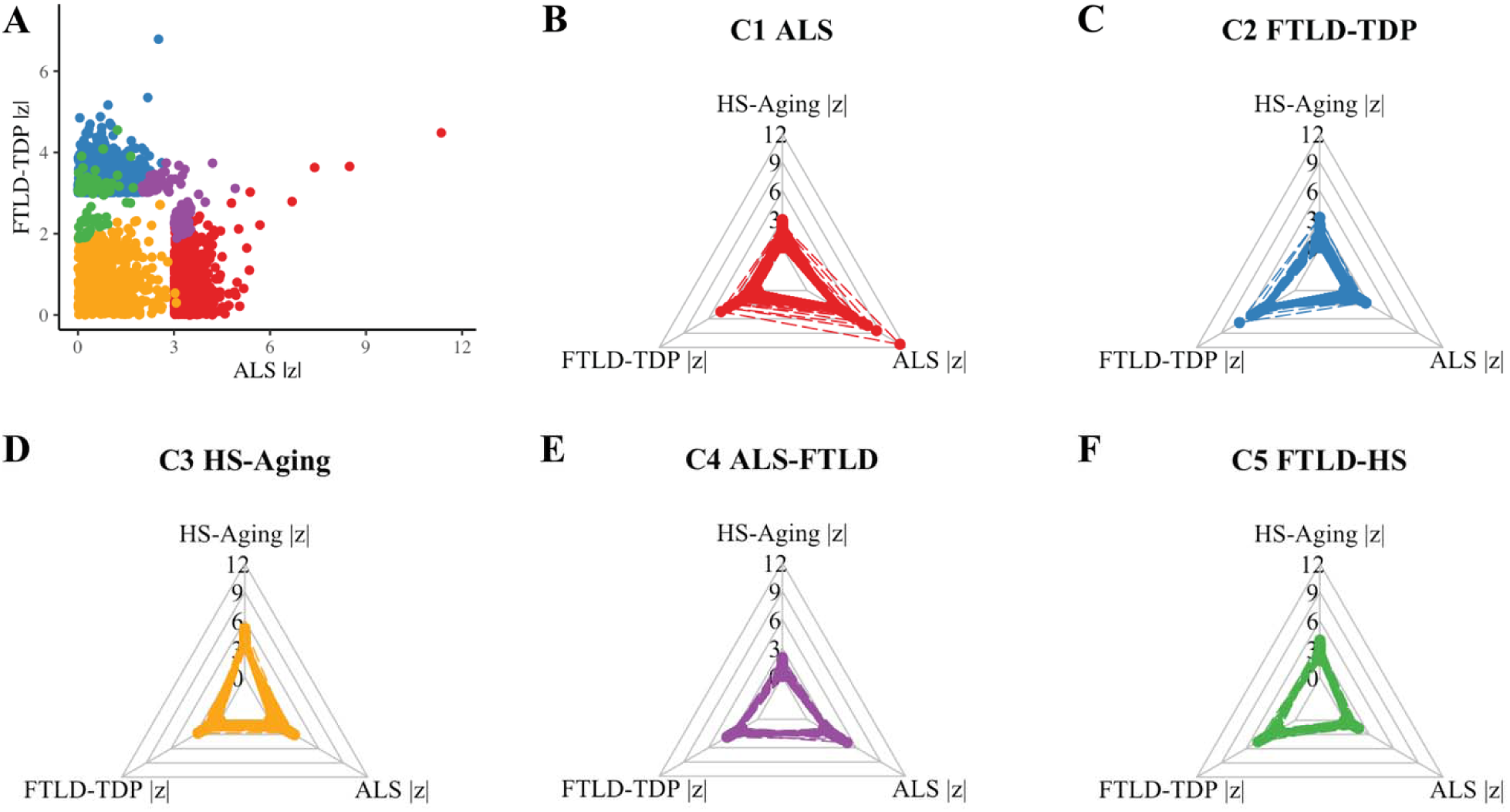
Weighted correlation analysis revealed clusters of correlated SNPs across GWAS phenotypes associated with TDP-43 proteinopathies. Cluster SNPs are plotted by relative associations (by |z|) for (A) all clusters, shown in two dimensions, FTLD-TDP and ALS, and for each cluster, (B) C1 ALS, (C) C2 FTLD-TDP, (D) C3 HS-Aging, (E) C4 ALS-FTLD, and (F) C5 FTLD-HS across FTLD-TDP, ALS, and HS-Aging GWAS.

### Pathway analysis revealed biologically plausible associations for cluster SNPs

Beyond individual SNPs, it is likely that biological pathways are involved in the development of complex neurodegenerative diseases. To investigate the biological pathway associations of each of our identified clusters, we performed a gene ontology enrichment analysis for each cluster using gprofiler2.^30^ Enrichment was determined by overrepresentation test, which calculates an expected value by matching the proportion of genes in a particular annotation data category in the human genome reference list to the number of genes provided. Cluster-specific overrepresented pathways suggest the identified clusters are biologically meaningful (FDR p<.05, Fig. 2 and Data file S1). Notably, C1 ALS uniquely included pathways related to motor neurons and spinal cord, while neurogenesis-related terms were among the top overrepresented pathways in C3 HS-Aging. Additionally, many pathways that were common across clusters related to nervous system processes that may be broadly implicated across neurodegenerative diseases (Fig. S4). Together, these pathway associations support the presence of differentiated and biologically meaningful genetic contributions within the identified clusters. Further, they reinforce the potential for shared susceptibility across diseases, while pointing to potential disparate drivers of ALS, FTLD-TDP, and HS-Aging.

**Fig. 2.**
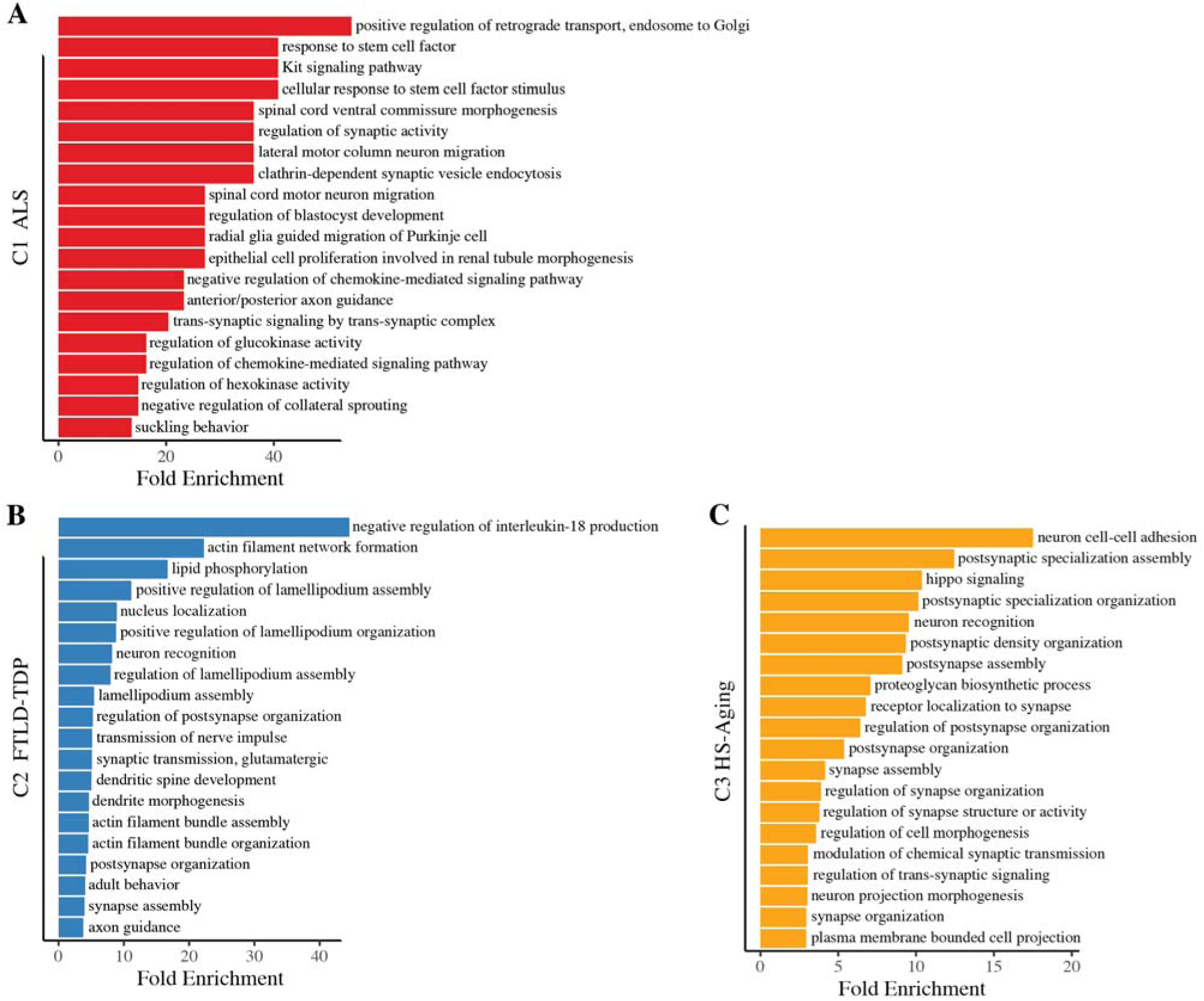
Derived clusters have biologically plausible pathway associations. The top 20 overrepresented pathways by fold enrichment that met FDR-adjusted p<.05 are displayed for (**A**) C1 ALS, (**B**) C2 FTLD-TDP, and (**C**) C3 HS-Aging.

### Cluster-specific polygenic risk associated with individual-level clinical and neuropathological trait heterogeneity

While the identified data-driven clusters suggest GWAS phenotype-specific SNPs which may contribute to our understanding of clinical and neuropathological trait heterogeneity across TDP-43 proteinopathies, these summary-level results alone cannot address individual-level risk. We focused on these GWAS phenotype-specific SNPs and hypothesized that they would contribute to individual-level differences in both clinical and neuropathological traits within the ALS-FTD clinicopathologic spectrum, where individuals have heterogeneous clinical, neuropathological, and genetic traits. Cluster-specific PRS were constructed for C1 ALS and C2 FTLD-TDP as the weighted sum of alleles per SNP per cluster for each individual, centered and mean scaled. The difference between an individual’s C2 FTLD-TDP risk and C1 ALS risk was evaluated to assess relative FTLD-TDP to ALS risk.

### Polygenic risk associates with clinical traits within mutation carriers at risk for neuromuscular and cognitive-behavioral impairment

To assess whether relative polygenic risk was associated with clinical trait heterogeneity across the ALS-FTD spectrum, the difference between C2 FTLD-TDP risk and C1 ALS risk was tested for association with clinical traits. A one-way ANOVA revealed that C2 FTLD-TDP risk, relative to C1 ALS risk, was higher in individuals with cognitive-behavioral impairment than those with MND, controlling for last contact age, sex, and inclusion in the prior ALS or FTLD-TDP GWAS (F(2,248)=[7.18], p<0.001), Fig. S5). Pairwise t-tests revealed an FDR-adjusted significant difference in the relative polygenic risk between the Cog-only and MND-only groups. This suggests that genetic variation associates with individual-level differences in clinical traits.

As our sample contained both sporadic and familial cases, polygenic risk was subsequently specifically tested for association with clinical trait heterogeneity within known *C9orf72*, *GRN*, *TARDBP*, *TBK1*, and *VCP* mutation carriers, where autosomal dominant inherited mutations may primarily drive disease more than underlying polygenic risk. As the Cog+MND group did not differ in relative risk from either the Cog-only or the MND-only group in the full sample, we tested cluster-specific polygenic risk against its corresponding trait, cognitive-behavioral impairment or neuromuscular impairment. One-way ANOVAs revealed that the C1 ALS PRS was higher in mutation carriers with MND (MND-only or Cog+MND, compared to Cog-only; F(1,125)=[6.87], p=0.01), while the C2 FTLD-TDP PRS was higher in mutation carriers with cognitive-behavioral impairment (Cog-only or Cog+MND, compared to MND-only; (F(1,125)=[12.47], p<0.001), controlling for last contact age, sex, and inclusion in the prior ALS or FTLD-TDP GWAS, respectively (Fig. S6). This suggests that even in the context of autosomal dominant disease, genetic variation associates with risk of a specific impairment including C1 ALS risk for neuromuscular impairment and C2 FTLD-TDP risk for cognitive-behavioral impairment.

Further limiting the sample to *C9orf72* cases which can develop ALS, FTLD-TDP, or both, a one-way ANOVA revealed that the C1 ALS PRS was higher in *C9orf72* carriers with MND (MND-only or Cog+MND, compared to Cog-only (F(1,104)=[4.2], p=0.04), while the C2 FTLD-TDP PRS was higher in *C9orf72* carriers with cognitive-behavioral impairment (Cog-only or Cog+MND, compared to MND-only; F(1,104)=[8.62], p=0.004), Fig. 3), controlling for last contact age, sex, and inclusion in the prior ALS or FTLD-TDP GWAS, respectively. This provides individual-level evidence that each cluster independently contributes to risk for a neuromuscular impairment or a cognitive-behavioral impairment in the context of a *C9orf72* expansion.

**Fig. 3.**
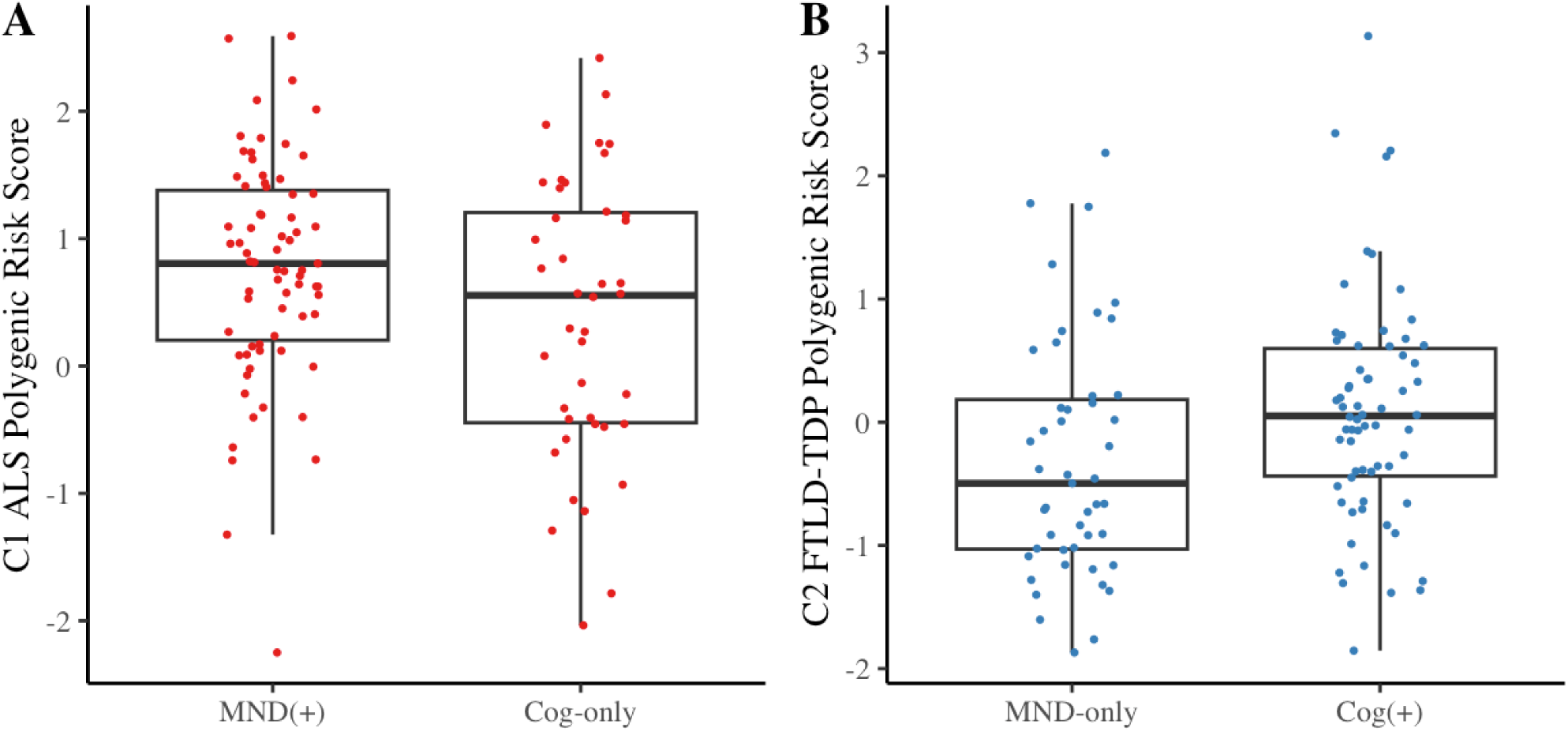
C1 ALS and C2 FTLD-TDP associate with individual-level clinical traits in C9orf72 cases. The (**A**) C1 ALS and (**B**) C2 FTLD-TDP PRS were tested for association with neuromuscular or cognitive-behavioral impairment or in C9orf72 mutation carriers.

### Polygenic risk associates with regional neuropathological traits

TDP-43 inclusions are observed in characteristic anatomic regions correlating with clinical traits. To assess whether relative polygenic risk was associated with regional neuropathological traits across the ALS-FTD spectrum, the difference between C2 FTLD-TDP risk and C1 ALS risk was tested for association with the burden of TDP-43 pathology across brain regions. First, this was done in characteristic FTLD-TDP regions, specifically the cingulate gyrus, angular gyrus, middle frontal cortex, and superior/middle temporal cortex (Fig. S7). One-way ANOVAs revealed that higher C2 FTLD-TDP risk, relative to C1 ALS risk, was associated with higher TDP-43 burden in each FTLD-TDP characteristic region (all FDR adjusted p < .05, Fig. 4), controlling for age at death, sex, and inclusion in the prior ALS or FTLD-TDP GWAS. Second, we tested whether relative polygenic risk was associated with TDP-43 burden in characteristic ALS regions, motor cortex and spinal cord. However, no significant differences in relative PRS by TDP-43 burden were observed in either ALS characteristic region. These observations suggest that these data-driven cluster-specific polygenic risk scores relate to characteristic individual-level differences in the anatomic distribution of TDP-43 within the ALS-FTD clinicopathologic spectrum.

**Fig. 4.**
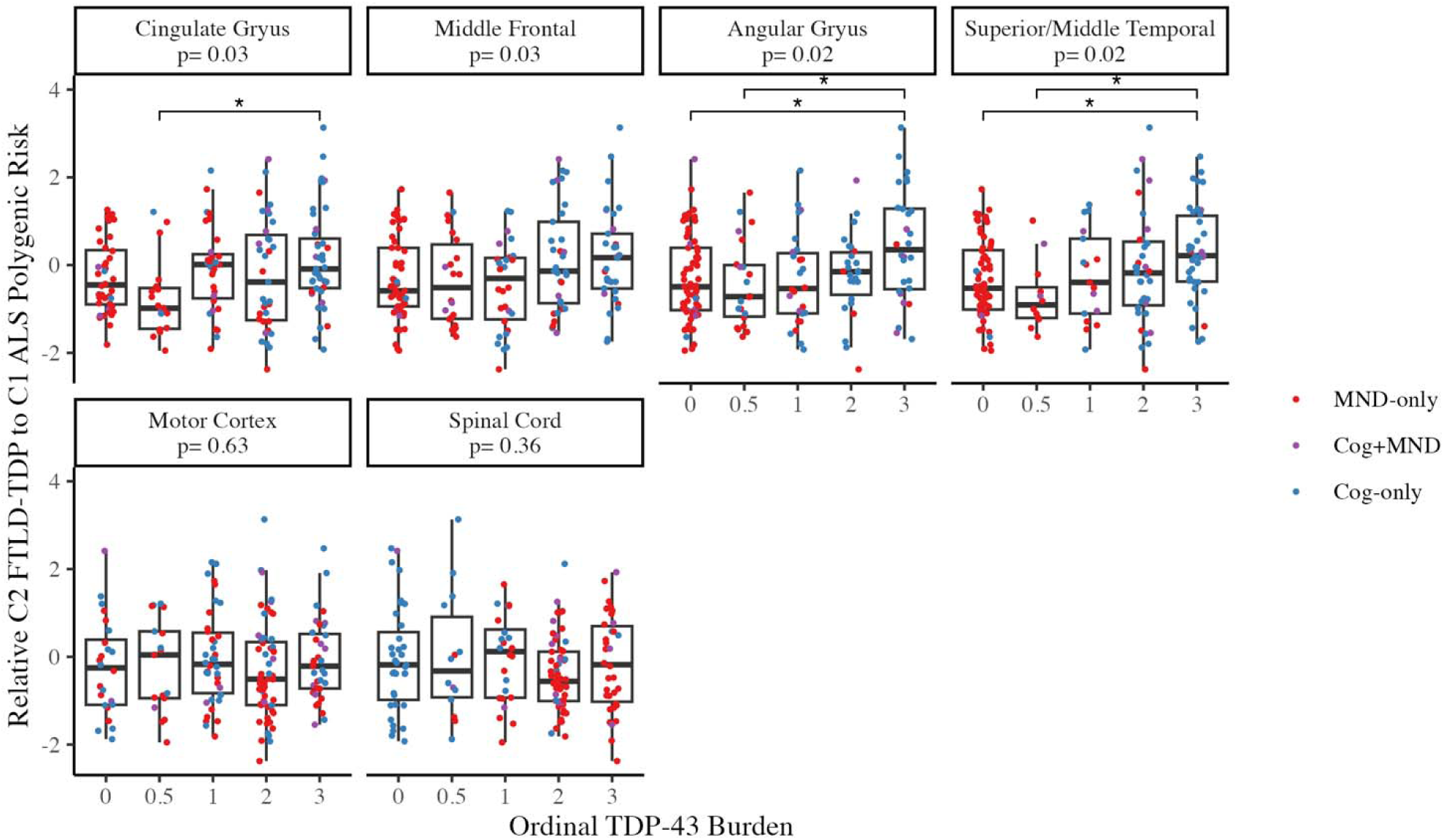
Relative polygenic risk associates with regional neuropathological traits. The difference between an individual’s C2 FTLD-TDP risk and C1 ALS risk was tested for association with the burden of TDP-43 pathology in FTLD-TDP characteristic regions (top row: cingulate gyrus, angular gyrus, middle frontal cortex, and superior/middle temporal cortex), and ALS characteristic regions (bottom row: motor cortex and spinal cord). The FDR adjusted p-values for each ANOVA are displayed by region. Asterisks represent FDR-adjusted significant pairwise t-tests within region.

## DISCUSSION

In this study, we hypothesized that both shared and disparate genetic variants would be identified across TDP-43 proteinopathies which contribute to the heterogenous individual-level clinical and neuropathological traits of ALS and FTLD-TDP. We identified data-driven clusters of correlated SNPs that were either GWAS phenotype specific (C1 ALS, C2 FTLD-TDP, and C3 HS-Aging) or shared across TDP-43 proteinopathy GWAS phenotypes (C4 ALS-FTLD and C5 FTLD-HS). Leveraging commonly genotyped SNPs across existing independent GWAS allowed for the identification of patterns of correlations between SNPs and the respective GWAS phenotypes. Though performed at the summary statistic level, this analysis suggested GWAS phenotype-specific cluster SNPs which were then shown to associate with clinical and neuropathological traits at the individual level, including neuromuscular impairment and/or cognitive-behavioral impairment and the burden of TDP-43 pathology across brain regions.

Gene ontology enrichment analysis of cluster SNPs provided plausible reinforcement for the presence of both shared and GWAS phenotype-specific biological pathways across TDP-43 proteinopathies. Terms relevant to nervous system development, such as neuron projection development and morphogenesis, as well as those related to synapse structure, organization, activity, and assembly were common across C1 ALS, C2 FTLD-TDP, and C3 HS-Aging, as would be expected in neurodegenerative diseases. However, top overrepresented pathways also support disease-specific drivers. Several of the top overrepresented pathways in C1 ALS related to the selective vulnerability of motor neurons and spinal cord in ALS. Among these were terms related to motor neuron migration, including spinal cord motor neuron migration and lateral motor column neuron migration. This analysis also identified several terms in C1 ALS related to axonogenesis, including central nervous system projection neuron axonogenesis and specifically spinal cord ventral commissure morphogenesis, as well as anterior/posterior axon guidance and negative regulation of collateral sprouting. Further, detailed molecular studies have identified inflammatory responses following neurodegeneration in ALS,^46,47^ and have implicated inflammatory pathways as contributors to further neurodegeneration.^48,49^ Gene expression studies in ALS have identified an upregulation of immune and inflammatory genes in disease relevant regions^50–53^. Here, we observed pathways related to the response to cytokines in C1 ALS, including gene ontology terms such as response to stem cell factor, cellular response to stem cell factor stimulus, cytokine-mediated signaling pathway, Kit signaling pathway, response to chemokine, and the regulation of chemokine-mediated signaling pathway. The top overrepresented pathway in C1 ALS was positive regulation of retrograde transport, endosome to Golgi. Golgi changes are well documented in ALS and one of the most consistent pathologic findings in motor neuron disease.^54–56^ Finally, C1 ALS included terms related to the regulation of metabolic process, including regulation of kinase activity, regulation of hexokinase activity, and regulation of glucokinase activity. Several lines of evidence support the role of kinases in ALS pathogenesis, including the identification of kinase-encoding genes such as *TBK1*, mutations in which can cause ALS.^57^ In C2 FTLD-TDP, several of the top overrepresented pathways related to cytoskeleton organization, including actin filament network formation, actin filament bundle organization, and actin filament bundle assembly. Differential expression of genes relating to the structure and function of the cytoskeleton have been observed in FTLD-TDP, and the cytoskeleton has been implicated more broadly in aging and neurodegeneration.^58–61^ For C3 HS-Aging, we observed several overrepresented terms related to neurogenesis and neuron development, including neuron recognition, which was among the top overrepresented pathways for this cluster. As the hippocampus is the site of adult neurogenesis, the inclusion of these pathways align with the selective vulnerability of the hippocampus in HS-aging.^62,63^

Within the ALS-FTD spectrum, the genetic architecture is complex. In addition to shared causal mutations, such as the *C9orf72* repeat expansion, shared significant associations have been identified in *UNC13A*.^20,23,24^ Similarly, *TMEM106B* has been implicated for risk in both FTLD-TDP and HS-Aging.^2,3,6^ Combined studies across TDP-43 proteinopathies have largely focused on identifying shared genetic risk factors that would explain susceptibility across GWAS phenotypes through a common underlying biological mechanism. In contrast, the identification of GWAS phenotype-specific cluster SNPs (e.g. C1 ALS and C2 FTLD-TDP), excluding those SNPs found to be shared across phenotypes (e.g. C4 ALS-FTLD), allowed us to demonstrate the contribution of genetic variants to individual-level clinical and neuropathological traits through the construction of cluster-specific PRS. Specifically, PRS analyses showed that higher C2 FTLD-TDP risk, relative to C1 ALS risk, was associated with higher TDP-43 burden in FTLD-TDP characteristic regions including cingulate gyrus, angular gyrus, middle frontal cortex, and superior/middle temporal cortex. Orbito-frontal regions are affected early in FTLD-TDP, even when the overall burden of pathology is lower, though more widespread cortical pathology is observed in advanced cases.^68^ In contrast, these regions are either spared or affected later in the course of ALS, where pathology spreads from motor neurons.^69^ While no significant differences in relative PRS by TDP-43 burden were observed in either ALS characteristic region evaluated, it is possible that this is confounded by neuron loss, particularly for the spinal cord where neuronal dropout can lower TDP-43 burden.

It is currently unknown why some individuals with TDP-43 pathology develop neuromuscular impairments while others develop cognitive-behavioral impairments. Both the C1 ALS PRS and the C2 FTLD-TDP PRS showed associations with their respective expected clinical traits, even within known mutation carriers where disease may be less influenced by underlying polygenic risk. Strikingly, even within individuals with a *C9orf72* repeat expansion, for which there is no current way to predict the development of ALS, FTLD-TDP, or both, the C1 ALS PRS and C2 FTLD-TDP PRS associated with individual-level clinical traits. While further validation is required, this observation has important implications for predictive genetic testing and management of clinical care. Moreover, current clinical trials focus on either ALS or FTLD-TDP populations in the absence of knowledge about individual risk. These novel PRS may therefore be important for prognostication and stratification in the context of interpreting outcomes in a combined trial.^64–67^

This study is limited by the complexity of diagnosis across TDP-43 proteinopathies. A more complete clinical evaluation of neuromuscular impairment in suspected FTLD-TDP cases and cognitive-behavioral impairment in ALS cases is needed to better characterize these individuals. Additional challenges to complete, accurate diagnoses in the FTD-ALS spectrum include a lack of TDP-43 biomarkers and often rapid progression which may preclude evaluation of the full clinical picture. Thus, there is a reliance on autopsy. However, the regional distribution of pathology alone cannot determine the presence of a secondary neuromuscular or cognitive-behavioral impairments within FTLD-TDP or ALS. Therefore, our individual-level analyses focused on clinical traits (neuromuscular impairment and/or cognitive-behavioral impairment) and neuropathological traits (burden of TDP-43 pathology across brain regions) rather than separate diagnostic groups.

Further, even though the vast majority of HS-Aging cases have TDP-43 pathology, the pathological definition does not require the presence TDP-43. The more recently defined Limbic-predominant age-related TDP-43 encephalopathy (LATE) relies only on the presence of TDP-43 in the amygdala and subsequently hippocampus and middle frontal gyrus.^70^ As hippocampal sclerosis is neither sufficient nor necessary for a LATE diagnosis, future work including a LATE GWAS is needed to disentangle the genotype associations with HS-aging from limbic TDP-43. In the present study we were unable to evaluate the relationship between C3 HS-Aging SNPs and individual-level differences in either clinical or neuropathological traits due to a lack of directionality information in the published GWAS summary statistics.

The threshold for inclusion of SNPs initially selected for analysis from the GWAS summary statistics was arbitrary. This threshold (the top 1% of SNPs with the strongest associations to any GWAS phenotype) is lenient by GWAS standards, including SNPs with summary statistic p-values as high as 0.003. However, strict GWAS correction would leave too few SNPs to compare across GWAS phenotypes, and sub-threshold associations are still likely informative. However, relaxing the threshold beyond a certain point would introduce non-meaningful SNPs which have no associations with any of the GWAS phenotypes. Importantly, we show that the SNPs chosen at this threshold are biologically meaningful with converging evidence from our summary statistic and individual level results.

Finally, similar to limitations of the vast majority of PRS studies, the cluster-specific polygenic scores were both predominantly derived from and tested on individuals of European ancestry. Therefore, future work is required to validate the broader applicability of these findings in a diverse population.

Overall, examining summary statistics revealed clusters of both shared and disparate correlated SNPs across GWAS phenotypes associated with TDP-43 proteinopathies. These clusters have biologically plausible pathway associations, and cluster SNPs relate to individual-level clinical and neuropathological traits providing suggestive evidence of a potentially predictive and prognostic genetic marker of neuromuscular and/or cognitive-behavioral impairment risk. We suggest that there are polygenic modifiers of clinical and neuropathological traits across TDP-43 proteinopathies that may contribute to individual-level differences, including the likelihood for developing FTLD or ALS.

## Acknowledgement statement

### Funding

National Institutes of Health grant F32AG079618 (BES), P01AG066597 (CTM), NS109260 (DJI), Penn Institute on Aging, and The DeCrane Family Fund for Primary Progressive Aphasia.

### Author contributions

Conceptualization: BES, CTM, DAW Methodology: BES, CTM, EBL

Investigation: BES, DJI, LE, CCQ, DA, MB, MG Visualization: BES, CTM

Funding acquisition: BES, CTM, DJI Project administration: CTM, DJI Supervision: CTM, DJI Writing—original draft: BES, CTM

Writing—review & editing: BES, DJI, VVD, ES, EBL, LE, CCQ, DA, MB, MG, DAW, CTM

### Competing interests

All authors declare they have no relevant competing interests.

### Data and materials availability

All summary statistic data was accessed from prior GWAS.^1–3^ Genotype, diagnosis, and neuropathological data may be requested and upon approval of reasonable requests may be shared with individual investigators. Data requests can be completed through the Penn Neurodegenerative Data Sharing Committee webform: https://www.pennbindlab.com/data-sharing

## Data Availability

All summary statistic data was accessed from prior GWAS. Genotype, diagnosis, and neuropathological data may be requested and upon approval of reasonable requests may be shared with individual investigators. Data requests can be completed through the Penn Neurodegenerative Data Sharing Committee webform: https://www.pennbindlab.com/data-sharing

## Notes

### Competing Interest Statement

The authors have declared no competing interest.

### Summary of Updates

Updated document for clarity throughout

